# Enhancing the Diagnostic Utility of ASL Imaging in Temporal Lobe Epilepsy through FlowGAN: An ASL to PET Image Translation Framework

**DOI:** 10.1101/2024.05.28.24308027

**Authors:** Alfredo Lucas, Chetan Vadali, Sofia Mouchtaris, T. Campbell Arnold, James J. Gugger, Catherine Kulick-Soper, Mariam Josyula, Nina Petillo, Sandhitsu Das, Jacob Dubroff, John A. Detre, Joel M. Stein, Kathryn A. Davis

## Abstract

**Background and Significance:** Positron Emission Tomography (PET) using fluorodeoxyglucose (FDG-PET) is a standard imaging modality for detecting areas of hypometabolism associated with the seizure onset zone (SOZ) in temporal lobe epilepsy (TLE). However, FDG-PET is costly and involves the use of a radioactive tracer. Arterial Spin Labeling (ASL) offers an MRI-based quantification of cerebral blood flow (CBF) that could also help localize the SOZ, but its performance in doing so, relative to FDG-PET, is limited. In this study, we seek to improve ASL’s diagnostic performance by developing a deep learning framework for synthesizing FDG-PET-like images from ASL and structural MRI inputs.

**Methods:** We included 68 epilepsy patients, out of which 36 had well lateralized TLE. We compared the coupling between FDG-PET and ASL CBF values in different brain regions, as well as the asymmetry of these values across the brain. We additionally assessed each modality’s ability to lateralize the SOZ across brain regions. Using our paired PET-ASL data, we developed FlowGAN, a generative adversarial neural network (GAN) that synthesizes PET-like images from ASL and T1-weighted MRI inputs. We tested our synthetic PET images against the actual PET images of subjects to assess their ability to reproduce clinically meaningful hypometabolism and asymmetries in TLE.

**Results:** We found variable coupling between PET and ASL CBF values across brain regions. PET and ASL had high coupling in neocortical temporal and frontal brain regions (Spearman’s *r >* 0.30, p < 0.05) but low coupling in mesial temporal structures (Spearman’s *r <* 0.30, p > 0.05). Both whole brain PET and ASL CBF asymmetry values provided good separability between left and right TLE subjects, but PET (AUC = 0.96, 95% CI: [0.88, 1.00]) outperformed ASL (AUC = 0.81; 95% CI: [0.65, 0.96]). FlowGAN-generated images demonstrated high structural similarity to actual PET images (SSIM = 0.85). Globally, asymmetry values were better correlated between synthetic PET and original PET than between ASL CBF and original PET, with a mean correlation increase of 0.15 (95% CI: [0.07, 0.24], *p*<0.001, Cohen’s *d* = 0.91). Furthermore, regions that had poor ASL-PET correlation (e.g. mesial temporal structures) showed the greatest improvement with synthetic PET images.

**Conclusions:** FlowGAN improves ASL’s diagnostic performance, generating synthetic PET images that closely mimic actual FDG-PET in depicting hypometabolism associated with TLE. This approach could improve non-invasive SOZ localization, offering a promising tool for epilepsy presurgical assessment. It potentially broadens the applicability of ASL in clinical practice and could reduce reliance on FDG-PET for epilepsy and other neurological disorders.

## Introduction

Temporal Lobe Epilepsy (TLE) is the most common type of epilepsy, presenting with seizures that can be localized to the temporal lobe via semiology and electroencephalogram (EEG) monitoring^1^. With nearly 1 in 3 patients with TLE having intractable seizures, surgical resection of the seizure-onset zone (SOZ) remains a common therapeutic intervention^2,3^. The success of surgical treatments, however, largely depends on accurate presurgical localization and lateralization of the SOZ. Neuroimaging techniques are essential during the presurgical assessment period. Current standard of care involves acquiring both MRI, to evaluate for causative structural lesions, and Positron Emission Tomography (PET) using fluorodeoxyglucose (FDG-PET), to detect areas of interictal hypometabolism associated with the SOZ^4,5^. However, despite its effectiveness, FDG-PET has limitations, including high cost and exposure to ionizing radiation, which may not be suitable for all patients^6,7^.

Arterial Spin Labeling (ASL) MRI offers a non-invasive and accessible complement to PET. ASL measures regional cerebral blood flow (CBF), which is normally coupled to regional cerebral metabolism^8^. ASL MRI demonstrates hypoperfusion (or hyperperfusion in certain instances) in regions associated with the SOZ^9–12^. Prior studies have compared the effectiveness of ASL hypoperfusion to that of PET hypometabolism as a biomarker for neurological disorders^13,14^, including epilepsy^15–18^. While some studies demonstrate comparable performance between PET and ASL for SOZ localization^15,16^, others show that PET still outperforms ASL^17^. An ability to approximate FDG-PET’s diagnostic performance using ASL, which can readily be acquired as part of a routine brain MRI protocol, would greatly extend the utility of MRI in the diagnosis and management of TLE.

To begin to address this need, our study introduces FlowGAN, a novel deep learning framework designed to enhance the diagnostic utility of ASL imaging in TLE, using the framework we established with LowGAN^19^, a low to high-field image translation model. FlowGAN improves on prior MR-to-PET algorithms^20,21^ by leveraging both perfusion information (ASL CBF maps) and structural information (T1-weighted MRI) to synthesize images that closely resemble those obtained from FDG-PET. By translating ASL and T1w inputs into PET-like images, FlowGAN provides a framework that could improve the clinical utility of ASL. This approach combines the accessibility and safety of ASL with the diagnostic superiority of PET, offering a promising tool for non-invasive and cost-effective localization of the SOZ in TLE.

In this study, we first validate the differences in regional coupling between metabolism and perfusion across brain regions and investigate how each can help lateralize the SOZ in epilepsy. We then introduce and test whether our image translation framework can successfully generate PET-like images from T1w and ASL inputs. Finally, we consider the feasibility of this image translation, and subsequently demonstrate the accuracy of FlowGAN-generated images in preserving and recovering critical hypometabolism asymmetries associated with TLE not originally apparent in the T1w and ASL inputs.

## Methods

### Study Design and Subject Selection

This study was designed as a retrospective analysis of imaging data from patients diagnosed with epilepsy from the Penn Epilepsy Center at the University of Pennsylvania. Inclusion criteria were: (1) a confirmed diagnosis of epilepsy, (2) availability of T1w and ASL MRI sequences, as well as FDG-PET, and (3) no history of neurological disorders other than epilepsy. The FDG-PET scans were acquired as part of the standard of care presurgical evaluation of each patient, whereas the ASL and T1w MRI sequences were from a research-specific protocol. We identified 68 subjects who fit these criteria with scan dates between June 2015 and January 2023. Demographic information is summarized in **Table 1**. Out of the 68 identified subjects, 36 were deemed to have lateralized TLE as per the surgical conference hypothesis. For the comparison of PET and ASL contrasts, only the 36 well lateralized TLE subjects were used. 19 of these well lateralized TLE subjects were selected as the test set for our deep learning architecture, FlowGAN, while the remaining 50 subjects of the entire epilepsy cohort were used as the training set. The Institutional Review Board of the University of Pennsylvania gave ethical approval for this work, and all subjects provided informed consent.

**Table 1.** Subject Demographics: Demographic and clinical characteristics for the entire cohort and the lateralized temporal lobe epilepsy only cohort. **Epilepsy location -** Lobar location of the seizure-onset zone. **MTS/HS** – Subjects with mesial temporal sclerosis/hippocampal sclerosis. **Laterality -** Seizure-onset zone lateralization as determined by surgical conference. **Surgical outcome –** Engel seizure outcome score. **TLE –** Temporal lobe epilepsy.

Subject Demographics
| Characteristic | Group |  |
| --- | --- | --- |
|  | All Epilepsy | Lateralized TLE |
| **Number of Subjects** | 68 | 36 |
| **Age** | 37±12 | 40±12 |
| **Gender (female)** | 35 | 20 |
| **Epilepsy location** | - |  |
| *Temporal* | 52 | 36 |
| *Frontal* | 4 | - |
| *Parietal* | 2 | - |
| *Other/Unknown* | 10 | - |
| **MTS/HS** | 19 | 12 |
| **Laterality** | - |  |
| *Left* | 33 | 23 |
| *Right* | 19 | 13 |
| *Bilateral* | 10 | - |
| *Unknown* | 5 | - |

### Imaging Data Acquisition

#### FDG-PET Imaging

Participants received interictal [F-18]FDG-PET/CT imaging as part of their presurgical evaluation. Scanning was performed on American College of Radiology accredited PET/CT instruments in accordance with Society of Nuclear Medicine and Molecular Imaging guideines^22^ (mean 10.3+/-0.7 mCi dose, mean uptake time 36.2 +/6.1 min, 10 minutes scanning duration).

#### MR Imaging

Arterial Spin Labeling (ASL) imaging was performed using a 3T Siemens PrismaFit scanner. The ASL sequence employed was a balanced 3D Pseudo-Continuous ASL (PCASL) with the following parameters: labeling duration of 1.8 seconds, post-labeling delay (PLD) of 1.8 seconds, 90% background suppression, and a 1D-accelerated single-shot stack-of-spirals readout with a nominal 3.75 mm isotropic resolution^23^. For quantitative analysis, the labeling efficiency was estimated at 0.72, and a separate M0 scan was acquired as a control for cerebral blood flow (CBF) quantification. High-resolution T1-weighted images, with a sagittal, 208-slice magnetization-prepared rapid gradient-echo (MP-RAGE) sequence, TE/TR = 2.24/2400 ms, with a 0.8 mm isotropic voxel size were also acquired in all subjects.

### Image Preprocessing

A detailed overview of the image preprocessing pipeline is shown in **Figure 1**. FreeSurfer was used to extract each subject’s Desikan-Killiany-Tourville (DKT) brain parcellation using the T1w images as input^24^. ASL data were preprocessed using ASLprep^25^ (**Supplementary Methods**), which performed motion correction, cerebral blood flow (CBF) estimation, and final registration to the native T1w images, which were also preprocessed with ASLprep. Computed CBF maps were then smoothed with a Gaussian kernel of σ=3 to enhance signal quality. For the FDG-PET data, we registered and resliced PET images to the native T1w images of each subject using ANTs affine registration^26,27^. Both PET and ASL images were visually inspected for quality control.

**Figure 1.**
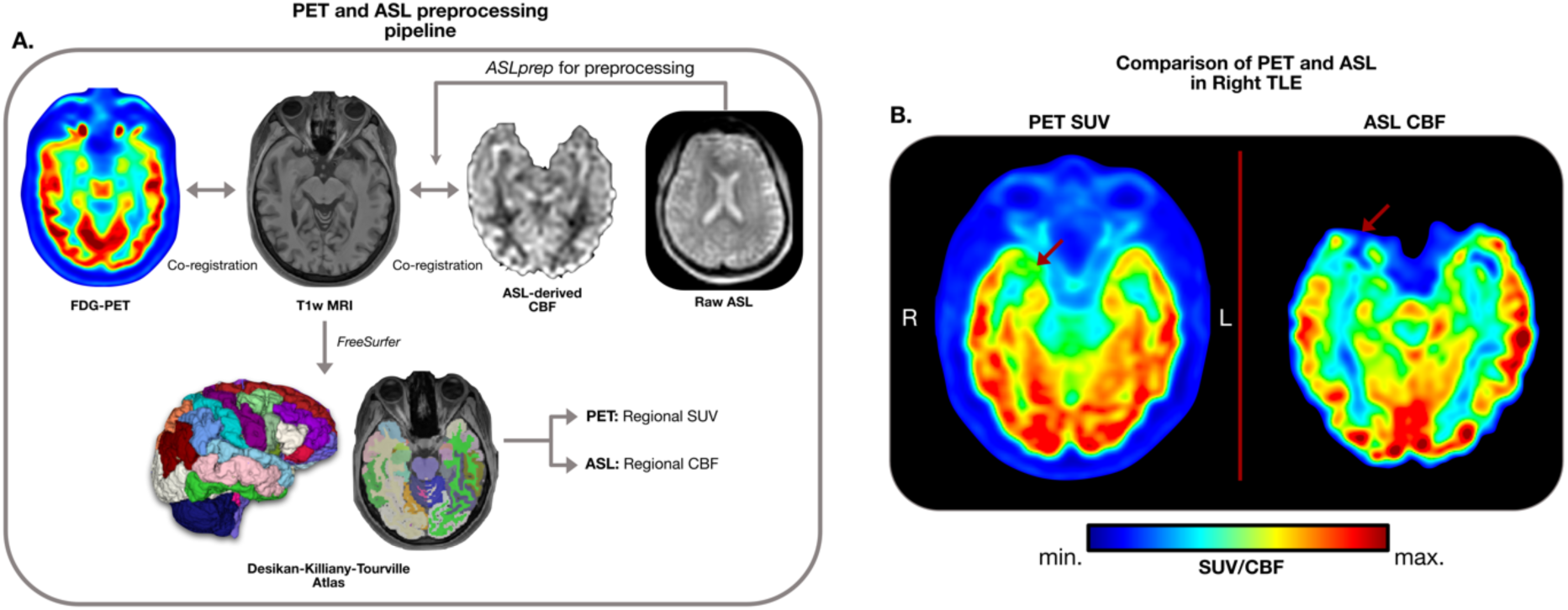
Image processing pipeline: **Panel A.** demonstrates the imaging pipeline used to pre-process the PET and ASL data for each subject. ASL data was preprocessed with ASLprep which performed cerebral blood flow (CBF) estimation and T1w MRI co-registration of the resulting CBF map. FDG-PET images were co-registered to the T1w image of each subject as well. Using FreeSurfer, Desikan-Killiany-Tourville atlas parcellations were extracted from each subject’s T1w image, and the corresponding PET SUV and ASL CBF values within each of these parcels was estimated. **Panel B.** shows the PET SUV and ASL CBF (Gaussian smoothed with σ=3) maps for the same subject. Red arrow points to a region of congruent hypometabolism and hypoperfusion.

### Regional PET SUVR and ASL CBF values

Using the subject-specific DKT parcellations, we extracted standardized uptake values (SUV) and CBF values from FDG-PET and ASL CBF maps respectively, for each cortical and subcortical brain region in the atlas. To do so, the mean contrast within each ROI was calculated. We generated FDG-PET standardized uptake value ratios (SUVR) by normalizing the ROI values, using the putamen as the control region. When comparing raw SUVR with CBF values (e.g. not asymmetry), the CBF values were also normalized relative to the putamen (rCBF)^14^.

### Regional PET SUV and ASL CBF asymmetry

In addition to raw SUVR and rCBF values, we also estimated the left-right asymmetries of these values, representing inter-hemispheric hypometabolism and hypoperfusion respectively, both of which are key biomarkers known to be associated with the SOZ. We estimated the asymmetry index (AI) between left and right brain regions as: AI = (left-right)/(left+right). Therefore, under this notation, a negative asymmetry in SUV/CBF would correspond to left sided hypometabolism/hypoperfusion, and a positive asymmetry would correspond to right sided hypometabolism/hypoperfusion.

### FlowGAN Training and Validation

We recently developed LowGAN, a low-field to high-field image translation generative adversarial network (GAN)^22^, which successfully generated 3T-like brain MRI images from T1w, T2w, and FLAIR inputs acquired on a 64mT scanner. Using this network architecture as a basis, we developed FlowGAN (**Figure 4**), a GAN for PET image synthesis from ASL and T1w inputs. FlowGAN consists of 3 parallel 2D pix2pix networks^23^, each of which processes input images from a different imaging plane (i.e. axial, coronal, and sagittal). Each parallel layer has three input channels, which consist of the T1w images and ASL-derived CBF maps smoothed with a Gaussian kernel of σ=1, and σ=3. The lower σ kernel is included to preserve finer perfusion features that might be removed by the larger kernel. The 2D outputs of each parallel pix2pix network are stacked into a 3D volume. In the original LowGAN implementation, a 3D U-Net was subsequently used to combine the 3 output volumes. However, because PET images are inherently smoother than structural images, we opted for using averaging and smoothing instead. First, the 3 output volumes were combined into a single volume by performing a voxel-wise averaging. Then, the final averaged image was smoothed using anisotropic diffusion with a conduction coefficient (kappa) of 80 and gamma of 0.1^24,25^, generating the final synthetic PET. To ensure that the final synthetic image was in the same space as the T1w inputs, a final rigid registration was applied to the model outputs, as small shifts are introduced by the architecture due to cropping and reshaping during model inference. As with the original PET images, the SUVR and asymmetry values across DKT brain regions were estimated using the FreeSurfer parcellations from the T1w inputs in the FlowGAN outputs.

The network was trained for 100 epochs with a batch size of 2, using the same set of hyperparameters as LowGAN^22^, on a subset of 50 epilepsy subjects. The trained model was then validated on a left-out subset of 19 subjects with well lateralized TLE.

### Statistical Analysis

Correlations were estimated with Spearman’s rank correlation coefficients, and comparisons between two metrics for the same subset of subjects were performed using paired two-tailed t-tests unless otherwise specified. Effect sizes were estimated using Cohen’s *d*, and statistical significance was defined at p<0.05. For all analyses, we adjusted the p-values for multiple comparisons across brain regions using the Benjamini-Hochberg False Discovery Rate (FDR) correction (pFDR).

#### Comparison of PET hypometabolism and ASL hypoperfusion in temporal lobe epilepsy

We analyzed the relationships between PET-derived SUVR, ASL-derived rCBF, and their asymmetries across various brain regions. To assess the coupling between metabolism and perfusion as measured with these two modalities, we calculated Spearman’s rank correlation coefficients to determine the statistical dependencies (coupling) between PET SUVR and ASL rCBF within each parcel of the Desikan-Killiany-Tourville (DKT) atlas. Additionally, we quantified the asymmetry index for PET SUV and ASL CBF across left and right brain regions, and subsequently computed correlations between these indices. We used the area under the ROC curve (ROC-AUC) to assess the capacity of PET SUV and ASL CBF asymmetries to lateralize TLE. Note that we did not create a classifier, but instead used the asymmetry values themselves as inputs to the ROC-AUC analysis. We calculated the ROC-AUC values for the brain regions in the DKT atlas, with higher values indicating enhanced discriminative ability between left and right TLE. Comparisons between ROC-AUC values, as well as estimation of their confidence interval, were done using DeLong’s test^26^. We also performed k-means clustering to classify patients based on regional asymmetry in PET SUV and ASL CBF across all brain regions combined, measuring the accuracy of features derived from each modality in distinguishing between left and right TLE. Finally, we conducted Principal Component Analysis (PCA) on the features from the k-means models, utilizing the first principal component for an additional ROC-AUC analysis to determine the separability of TLE types across all brain features.

#### Assessment of FlowGAN outputs

We employed the Structural Similarity Index (SSIM) to compare synthetic PET images generated by FlowGAN with actual PET images. Furthermore, we calculated the Spearman correlation between original and synthetic PET images, as well as between original PET and ASL CBF images, across DKT brain regions to evaluate the fidelity of FlowGAN in replicating PET image patterns. We also introduced a congruency metric to assess the matching of hypometabolism/hypoperfusion patterns between PET, synthetic PET and ASL modalities. This metric calculated the proportion of samples exhibiting matching asymmetry between the two modalities.

## Results

### The coupling between PET SUVR and ASL CBF differs across brain regions

Using the subject specific Desikan-Killiany-Tourville (DKT) atlas parcellations derived from FreeSurfer, we quantified the PET-derived SUVR and the ASL-derived rCBF (**Figure 1**) for each subject. We found that the coupling between metabolism (SUVR) and perfusion (rCBF) across subjects in each DKT parcel was highly variable (**Figure 2A**, **Supplementary Table 1**). The highest coupling was found in the left superior parietal gyrus (Spearman’s *r* = 0.50, *p_FDR_* < 0.001), but most of the remaining regions of significant coupling were found in the right hemisphere, including the right inferior gyrus (Spearman’s *r* = 0.32, *p_FDR_* = 0.033) and the middle temporal gyrus (Spearman’s *r* = 0.32, *p_FDR_* = 0.033), as well as the right rostral gyrus (Spearman’s *r* = 0.45, *p_FDR_* = 0.003) and the caudal middle frontal gyrus (Spearman’s *r* = 0.41, *p_FDR_* = 0.006). Except for the right caudate and accumbens area, no other subcortical structure had a significant coupling between CBF and SUVR. Notably, the hippocampi (left hippocampus: Spearman’s *r* = -0.14, *p_FDR_* = 0.380, right hippocampus: Spearman’s *r* = 0.08, *p_FDR_* = 0.610) and the amygdalae (left amygdala: Spearman’s *r* = -0.01, *p_FDR_* = 0.920, right amygdala: Spearman’s *r* = 0.04, *p_FDR_* = 0.817), key structures for the diagnosis of epilepsy, had very low coupling values. A sorted list of correlation values across brain structures is presented in **Supplementary Table 1**. These findings demonstrate a regionally dependent pattern of coupling between PET SUVR and ASL rCBF, with high coupling in frontoparietal and neocortical temporal lobe regions and notably low coupling in mesial temporal structures.

**Figure 2.**
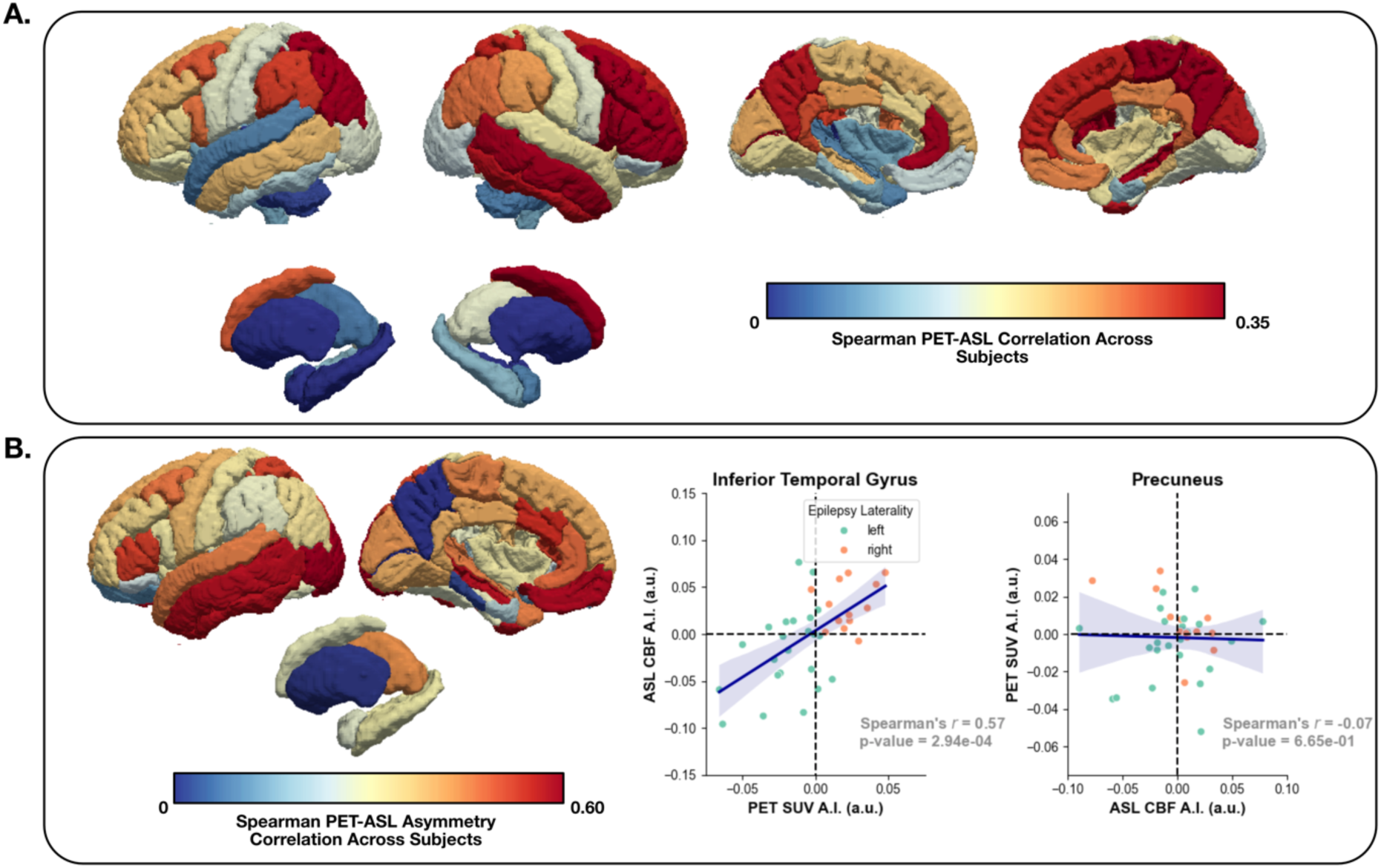
SUVR and CBF raw and asymmetry regional correlations: **Panel A.** shows the Spearman rank correlation between PET SUVR and ASL CBF across subjects for each DKT brain region. **Panel B.** (left) shows the Spearman rank correlation between PET SUV and ASL CBF left-right asymmetry across subjects for each DKT brain region; (middle) shows the asymmetry scatterplot between PET and ASL asymmetry values for the inferior temporal gyrus parcel (high correlation between PET and ASL); (right) shows the asymmetry scatterplot between PET and ASL asymmetry values for the precuneus parcel (low correlation between PET and ASL). In the scatterplots, each dot represents a subject, and left TLE and right TLE subjects are shown in different colors. Dashed lines represent zero asymmetry.

### Asymmetry values are better coupled between PET and ASL

We quantified the asymmetry index between left and right DKT parcels for both PET SUV and ASL CBF. We then determined the correlation between this asymmetry measured for each modality (**Figure 2B**; **Supplementary Table 2**). Overall, the asymmetry correlations across regions were larger than the correlations for SUVR and rCBF values (asymmetry correlations: 0.33±0.18, 95% CI: [0.27, 0.39]; SUVR and rCBF correlations: 0.18±0.13, 95% CI: [0.15, 0.21]; p<0.001, Cohen’s *d* = 0.96), with the largest correlation in the lateral occipital gyrus (Spearman’s *r* = 0.64, *p_FDR_* <0.001) followed by the medial orbitofrontal cortex (Spearman’s *r* = 0.57, *p_FDR_* = 0.002), and the middle (Spearman’s *r* = 0.57, *p_FDR_* = 0.002) and inferior temporal gyri (Spearman’s *r* = 0.57, *p_FDR_* = 0.002). The other significant correlations were located predominantly in the temporal and frontal lobes. For subcortical structures, the asymmetry correlation was higher than for raw SUVR and rCBF values, but the thalamus was the only subcortical structure with a significant correlation (Spearman’s *r* = 0.43, *p_FDR_* = 0.02). A sorted list of asymmetry correlation values is presented in **Supplementary Table 2**. These findings demonstrate a higher coupling between the left-right asymmetry of CBF and SUV asymmetry than between raw SUVR and rCBF values. This is relevant for lateralized epilepsy, as the asymmetry between left hemisphere and right hemisphere PET contrast is commonly used for non-invasively lateralizing the SOZ^4,5,28–30^, and these findings demonstrate that in certain brain regions, ASL-derived CBF can approximate this asymmetry.

### Both PET and ASL quantified asymmetries can lateralize TLE

We quantified whether SUV and CBF asymmetries could distinguish left and right TLE across brain regions. We used the area under the receiver operating characteristics curve (ROC-AUC) of the regional asymmetry as a measure of separability between left and right TLE, with larger ROC-AUC values corresponding to better separability. We found that asymmetries across brain regions have different abilities to lateralize TLE depending on whether PET or ASL is used. For PET SUV asymmetry, the top 2 performing regions were the inferior temporal gyrus (AUC = 0.97, 95% CI: [0.90, 1.00]), and the middle temporal gyrus (AUC = 0.95, 95% CI: [0.85, 1.00]), for which the ASL CBF asymmetry also had high AUCs (inferior temporal gyrus: AUC = 0.83, 95% CI: [0.67, 0.98]; inferior temporal gyrus: AUC = 0.96, 95% CI: [0.71, 0.99]) (**Figure 3A**). As for the mesial temporal structures, both the amygdala and hippocampus had high lateralizing ability for PET SUV asymmetries (amygdala: AUC = 0.87, 95% CI: [0.73, 1.00]; hippocampus: AUC = 0.93, 95% CI: [0.82, 1.00]), but not for ASL CBF (amygdala: AUC = 0.55, 95% CI: [0.34, 0.74]; hippocampus: AUC = 0.70, 95% CI: [0.51, 0.88]). In fact, the largest difference in AUCs between modalities **(Figure 3C)** was in the amygdala (AUC difference = 0.33, DeLong’s test *p_FDR_* = 0.019). A list of TLE lateralization AUCs for PET and ASL derived asymmetries, as well as their difference, is presented in **Supplementary Table 3**. These findings demonstrate comparable lateralization performance in neocortical temporal regions between PET and ASL asymmetries, but this performance drops significantly for ASL in mesial temporal structures.

**Figure 3.**
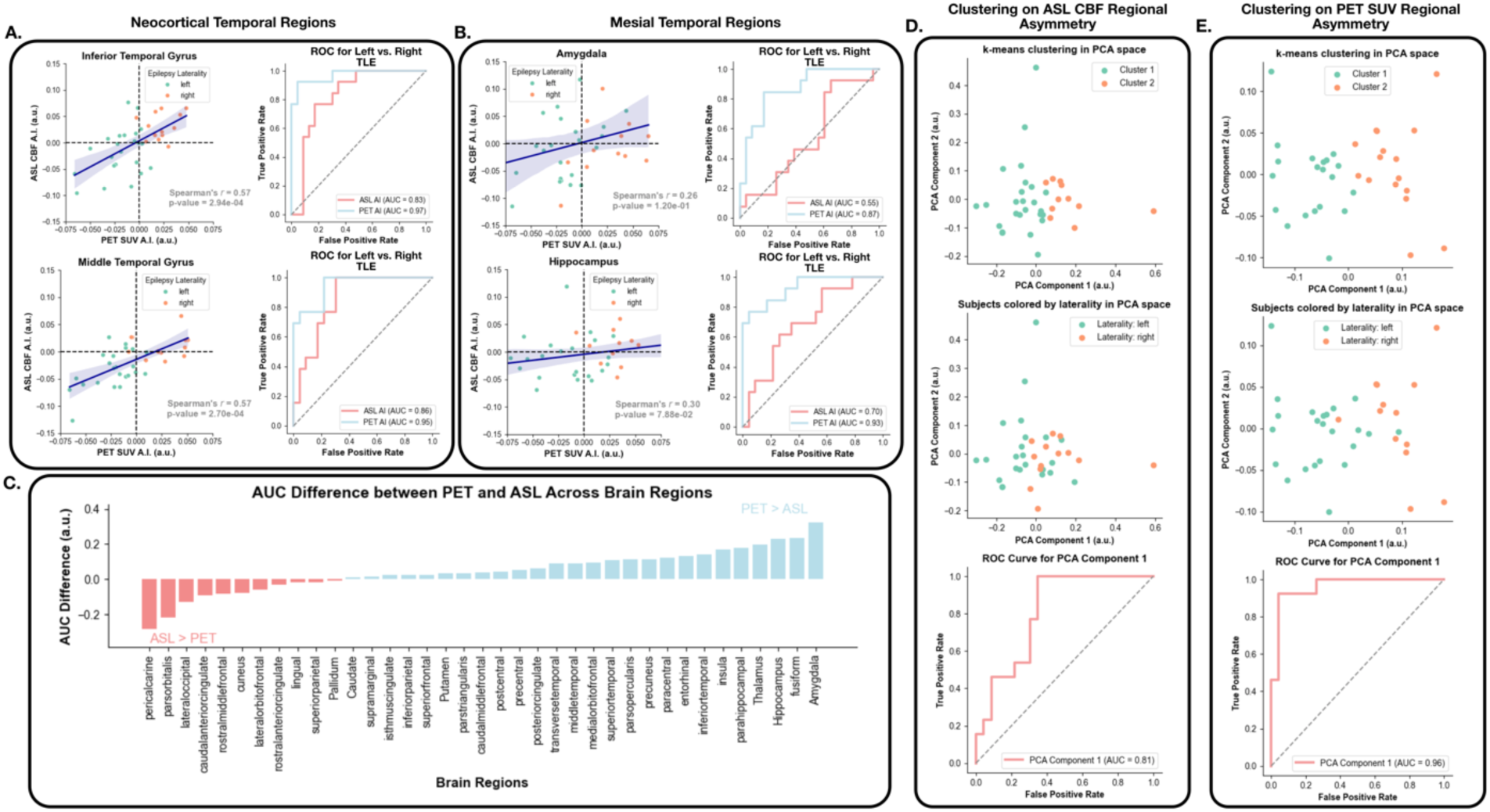
Comparison of SOZ lateralization between PET and ASL: **Panel A.** shows the asymmetry scatterplot between PET and ASL asymmetry values as well as the corresponding ROC for separating left TLE from right TLE using PET or ASL asymmetry in the (top) inferior temporal gyrus and (bottom) middle temporal gyrus. **Panel B.** shows the same but in the amygdala (top) and hippocampus (bottom). In the scatterplots, each dot represents a subject, and left TLE and right TLE subjects are shown in different colors. Dashed lines represent zero asymmetry. **Panel C.** shows the difference in the area under the ROC curve between PET and ASL asymmetry across brain regions. **Panel D.** shows a scatterplot of the first two principal components generated by the ASL CBF asymmetry values across all brain regions, with (top) each subject colored according to their cluster assignment after k-means clustering, and (middle) each subject colored according to their epilepsy laterality. The ROC curve at the bottom shows the separability between left and right TLE based on the values of the first principal component. **Panel E.** shows the same as **D.** but for the PET SUV asymmetry values. **A.I.** asymmetry index.

We also clustered the subjects using the asymmetry across all brain regions as features in the same model. We created two k-means clustering models, one that clustered patients based on ASL CBF regional asymmetry (**Figure 3D**) and one that clustered patients based on PET SUV regional asymmetry (**Figure 3E**). The accuracy of the ASL-based asymmetry clustering was 66% and that of the PET-based asymmetry classifier was 86%. We then performed principal component analysis on the features of both classifiers and used the first principal component as an input to a ROC-AUC analysis and found that the separability of left and right TLE in the space of the first principal component was high for both the PET and the ASL features. Separability was higher for the PET features (AUC = 0.96, 95% CI: [0.88, 1.00]) than for the ASL features (AUC = 0.81; 95% CI: [0.65, 0.96]), but the difference between the two was not statistically significant (AUC difference = 0.15, DeLong’s test *p* = 0.067). Overall, PET provides a better avenue for lateralizing the SOZ than ASL does, however, ASL CBF hypoperfusion is still capable of lateralizing TLE.

### FlowGAN successfully generates PET-like images from ASL and T1w inputs

The results describing FlowGAN outputs correspond to the model inference run on the held-out test-set of 19 subjects with well lateralized TLE. Visually, the resulting synthetic PET images are very similar to the actual PET images (**Figure 4**). As seen in **Figure 5A** and **Figure 5B**, the FlowGAN output reproduces the hypometabolism, even when the input T1w and CBF maps do not have clear visual abnormality. Quantitatively, the structural similarity (SSIM) between FlowGAN outputs and actual PET images for the test set was 0.85 (95% CI: [0.83, 0.87]), whereas between ASL CBF maps and actual PET images it was significantly lower at 0.26 (95% CI: [0.21, 0.31]), with a difference in SSIM of 0.59 (95% CI: [0.54, 0.64]; *p* < 0.001). We additionally quantified the regional correlation between the raw synthetic PET SUVR and original PET SUVR contrasts and compared them to the correlation between the raw original PET SUVR and ASL rCBF contrasts (**Figure 6A**). The average Spearman correlation across DKT brain regions for all test set subjects between original and synthetic PET SUVR was 0.89 (95% CI: [0.86, 0.92]), significantly higher than between original PET SUVR and ASL rCBF, where it was 0.57 (95% CI: [0.50, 0.64]), with a correlation difference between the two of 0.32 (95% CI: [0.25, 0.38], *p*<0.001, Cohen’s *d* = 2.29). These findings demonstrate that FlowGAN generates outputs images that are PET-like, and that preserve the general patterns of PET contrast across the brain.

**Figure 4.**
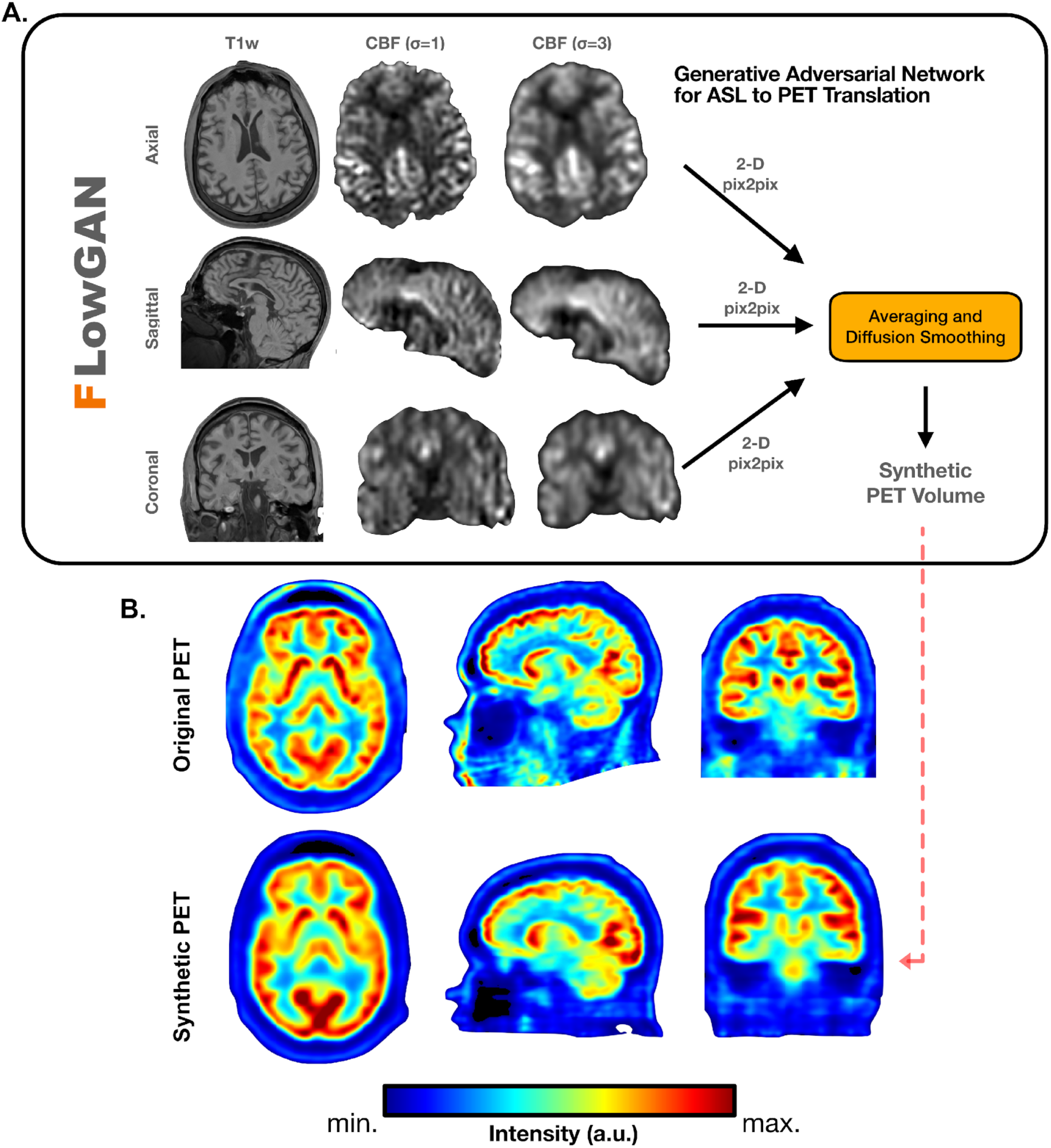
FlowGAN overview and representative test subject output: **Panel A.** shows an overview of the FlowGAN architecture, with axial, sagittal, and coronal inputs into 3 parallel pix2pix networks that are eventually combined through averaging and diffusion smoothing. **Panel B.** shows the original PET as well as the corresponding synthetic PET across all three imaging planes for the same subject.

**Figure 5.**
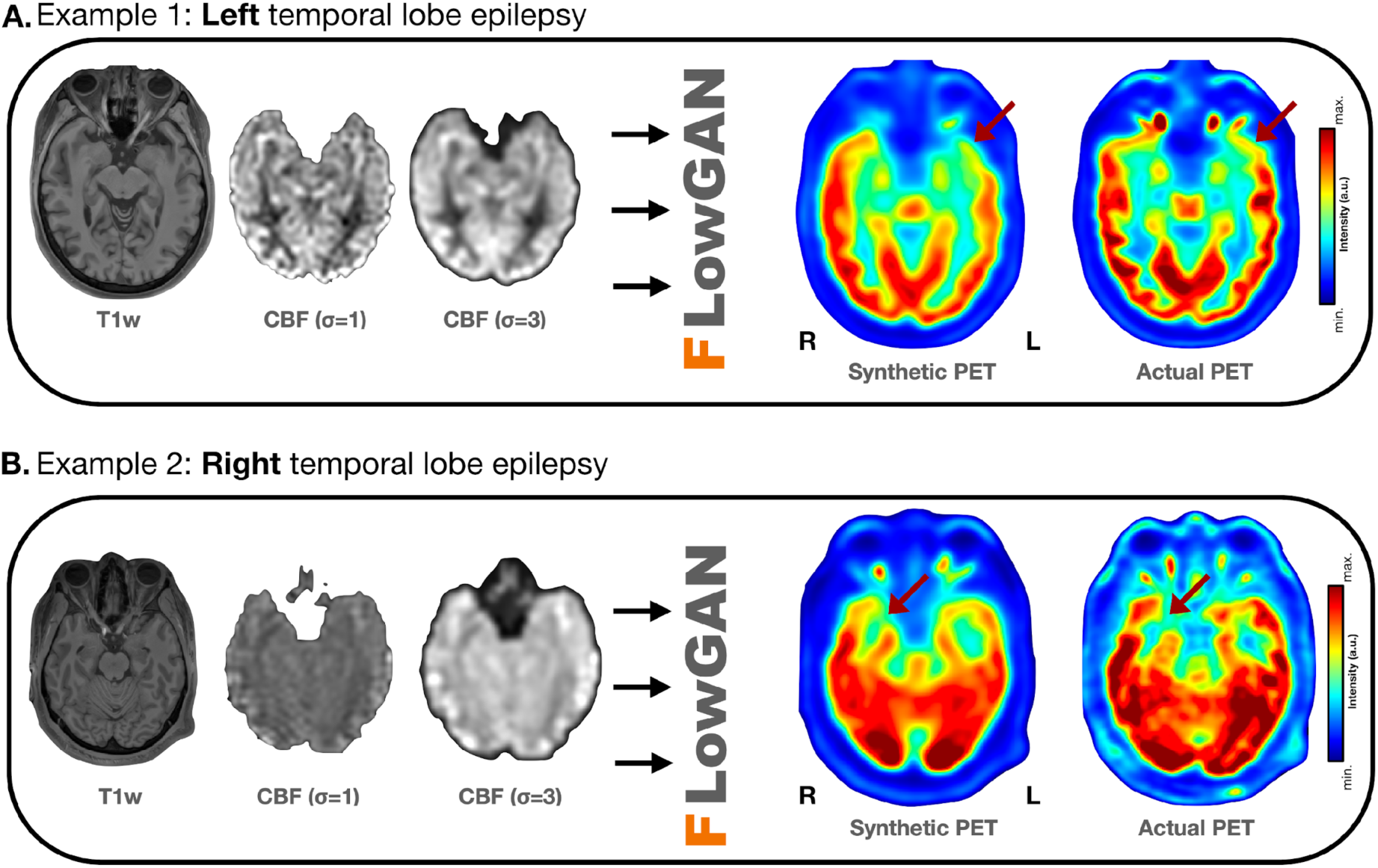
FlowGAN outputs for left and right TLE example test set subjects: Three co-registered inputs to FlowGAN as well as the corresponding synthetic PET output, and the actual PET image for a left TLE (**A.**) and a right TLE (**B.**) subject. In both cases, there is a region of hypometabolism in the temporal lobe (red arrow) consistent with the lateralization of the SOZ in both the synthetic and actual PET images. Both subjects were left-out subjects not seen by the model during training.

**Figure 6.**
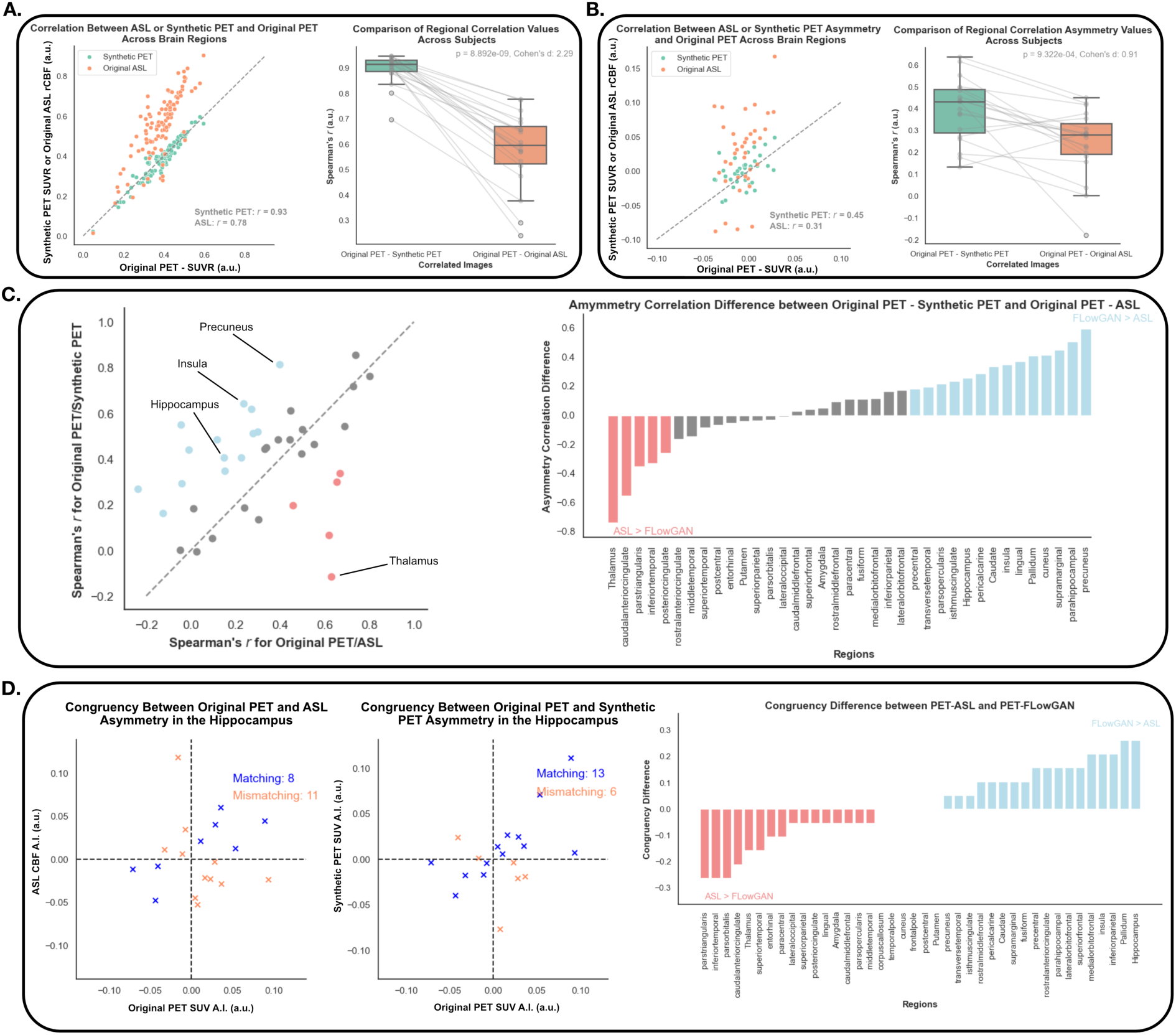
Relationship between FlowGAN outputs and original PET images across brain regions: **Panel A.** (left) shows a representative scatterplot between synthetic PET and original PET (green) values as well as original ASL CBF and original PET (orange) values for a single subject across brain regions. Each point represents one brain region, and the dashed line represents the unity line. The right side shows the boxplot comparing the correlations for all subjects across brain regions for original PET and synthetic PET for one boxplot, and original PET and original ASL for the other boxplot. The same subjects in each boxplot are connected by a line. **Panel B.** shows the same as **A.** but for asymmetry values across brain regions. **Panel C.** (left) shows the scatterplot between original PET - synthetic PET, and original PET – original ASL CBF asymmetry correlations across subjects for the different brain regions. Each dot represents a different brain region. Blue dots show an asymmetry correlation difference in favor of synthetic PET of more than 1 standard deviation, and red dots represent an asymmetry correlation difference in favor of ASL of more than 1 standard deviation. Gray dots are under the 1 standard deviation threshold. The right side of **C.** shows the asymmetry correlation difference between the two approaches across brain regions. **Panel D.** shows the scatterplot between (left) original ASL CBF and PET asymmetry values and (middle) synthetic PET and original PET asymmetry values across subjects in the hippocampus. In both cases, subjects that have the same direction of asymmetry (i.e. both positive, or both negative) between the two compared modalities (i.e. subjects with congruent measurements) are shown as blue **Xs**, whereas subjects with incongruent measurements are shown as red **Xs**. The right side shows the congruency difference between the two approaches across brain regions.

### FlowGAN recovers hypometabolism in regions with low PET-ASL coupling

We hypothesized that FlowGAN could help recover asymmetries in metabolism in brain regions where ASL alone is not capable of detecting asymmetries compared to PET, as demonstrated in the previous sections. We compared the Spearman correlation of asymmetry indices across brain regions between original PET SUV and synthetic PET SUV, as well as between original PET SUV and ASL CBF (**Figure 6B**). As with raw contrast correlations, the regional asymmetry correlations between original and synthetic PET were significantly higher (0.39, 95% CI: [0.32, 0.46]), than between original PET and ASL (0.24, 95% CI: [0.16, 0.31]), with a mean correlation difference of 0.15 (95% CI: [0.07, 0.24], *p*<0.001, Cohen’s *d* = 0.91). Out of the 19 test set subjects, the correlation between original and synthetic PET was higher in 17. These findings are evidence that generally across brain regions, FlowGAN can improve the correlation between metabolism and perfusion left-right asymmetry originally present between PET and ASL. Looking at the correlation in asymmetry across subjects for any given region, we can better understand the regions where FlowGAN provides the greatest benefit relative to ASL (**Figure 6C**). Comparing the asymmetry correlation across subjects each region, we find that FlowGAN has higher correlation (defined as a correlation higher than 1 standard deviation of the mean difference in correlations across all regions between original PET-synthetic PET and original PET-ASL) in 14/37 regions, whereas ASL has higher correlation in 5/37 regions, with no improvement above 1 standard deviation for 18/37 regions. Notably, several mesial temporal structures which we demonstrated had poor coupling between PET and ASL, now have significantly improved coupling. For example, the hippocampus (correlation improvement = 0.27), insula (correlation improvement = 0.34) and parahippocampal gyrus (correlation improvement = 0.50), all had improved correlations. Among the few correlations that decreased in the synthetic PET relative to the ASL, we have the thalamus (correlation decrease = -0.74). A list of asymmetry correlation values between original PET and ASL, original and synthetic PET, and their difference, for the test set subjects is presented in **Supplementary Table 4**.

In addition to comparing the asymmetry correlations, we also compared the matching of hypometabolism/hypoperfusion between modalities. For this analysis, if one modality had a positive asymmetry (left>right) and the other modality also had a positive asymmetry, that is considered a match between modalities for a given subject. Similarly for negative hypometabolism. We defined the congruency between the two modalities as the number of samples with matching positive or negative asymmetry, divided by the total number of samples. This is different from comparing asymmetry correlations, as two brain regions can be correlated, but still have mismatches in asymmetry due to variance, and this comparison better approximates what is done in clinical practice. As an example (**Figure 6D**), we show how the congruency increases in the hippocampus when synthetic PET is used, compared to when ASL is used, going from 42% congruency to 68% congruency with the original PET hypometabolism. This improvement in congruency, as with the correlation, was primarily in regions where the original ASL and PET images had poor correlation such as mesial temporal structures (**Figure 6D; Supplementary Table 5**).

## Discussion

In this study we demonstrate that FDG-PET metabolism and ASL CBF have a distinct pattern of coupling across cortical and subcortical brain regions. This coupling, both of raw values as well as left-right asymmetries, appears stronger in the neocortical temporal and frontal regions, with a much lower coupling in subcortical structures, where FDG-PET exhibits strong hypometabolism ipsilateral to the SOZ. These differences in coupling across key epileptogenic regions, like the mesial temporal structures, limit the applicability of ASL hypoperfusion as a biomarker for localization of the SOZ. As a solution, we introduced FlowGAN, an image translation framework that generates PET-like images from T1w and ASL CBF maps. Our study demonstrates that in regions where ASL was previously incapable of recovering meaningful asymmetries (e.g. the hippocampus), FlowGAN outputs provide meaningful asymmetries that are consistent with the original PET images.

The differences between ASL and PET coupling across the brain could be attributed to several factors. First, it is possible that physiologic neurovascular coupling differences in these brain regions lead to increases in brain perfusion that are not matched by concordant increases in metabolism^31^. Second, this analysis was done in subjects with epilepsy, which might have aberrant neurovascular coupling in the first place^32–34^. Our population, which was biased towards left-sided TLE, mostly showed significant coupling in the right hemisphere, potentially suggesting a pathological decrease in neurovascular coupling in the left hemisphere and subcortical regions due to influence from the SOZ. It is possible that this decrease in neurovascular coupling can itself be a marker of epileptogenicity, and future work could emphasize studying this. A third and final reason for the low PET-ASL coupling could be due to limitations in the ASL acquisition protocol. In particular, the lengthy single-shot readout used to acquire the ASL images causes considerable blurring in the slice direction (superior-inferior axis) that reduces CBF accuracy in small regions. Additionally, the ASL protocol used balanced pseudocontinuous labeling, which is less robust to magnetic field inhomogeneities than the unbalanced approach^35^.

Our results demonstrate that FlowGAN synthetic PET outputs not only accurately reproduce whole brain patterns of PET contrast, but also improve the hypometabolism correlation in regions where ASL had low coupling. It is possible that this improvement results from the T1w images providing additional structural information that ASL cannot resolve. Because metabolism is a function of the amount of neural tissue and blood flow, the addition of morphometric data may have allowed FlowGAN to more closely approximate PET in areas with poorer ASL resolution. Importantly, FlowGAN was able to preserve the general patterns across the brain and our biomarker of interest through the image translation process. Our results have implications beyond epilepsy and could suggest utility in disorders such as Alzheimer’s disease and other forms of dementia^13^. Future work will assess whether transfer learning, by training FlowGAN with an epilepsy cohort and then testing on MCI and Alzheimer’s disease subjects (and vice-versa), allows preservation of the FDG-PET hypometabolism commonly seen in these neurological conditions.

The results from this study also serve as validation of the generalizability of the image translation architecture proposed by our prior LowGAN work^19^. While the original architecture was developed for low-field to high-field brain MRI translation, the results of our FlowGAN implementation demonstrate successful image translation in a different domain (i.e. PET synthesis). This supports the notion that this image translation framework is readily adaptable for different medical imaging tasks.

Our study has certain limitations. In this proof-of-concept work, we demonstrate that FlowGAN can recover disease-relevant hypometabolism from ASL and T1w inputs. However, this does not directly prove the clinical utility of our approach. Future work could compare traditional PET imaging and FlowGAN-enhanced ASL imaging during presurgical evaluation for TLE, ideally using an updated ASL protocol with unbalanced labeling and a multi-shot readout, with the goal of demonstrating that synthesized PET-like images can effectively localize the SOZ. Successful validation of the diagnostic potential of FlowGAN-enhanced ASL would provide an alternative modality to FDG-PET.

## Conclusion

FlowGAN brings ASL closer to FDG-PET’s diagnostic performance, generating synthetic PET images that mimic actual PET in depicting hypometabolism associated with TLE. This approach could improve the non-invasive localization of the SOZ, offering a promising tool for presurgical assessment of epilepsy patients and potentially broadening the applicability of ASL in clinical practice.

## Data Availability Statement

Upon publication of our manuscript, we will release our GitHub repository with code to train FlowGAN, as well as the pre-trained weights used in this manuscript, such that other users can readily perform image translation from ASL and T1w to FDG-PET. We will also release the code used to generate the figures and the analysis described in this manuscript.

## Acknowledgments

AL and KAD received support from NINDS (R01NS116504). JD received support from NIDA (K23DA038726). JAD received support from NIBIB (P41EB029460).

## Author Contributions

**Alfredo Lucas**: Conceptualization, Methodology, Software, Validation, Formal analysis, Writing - Original draft preparation. **Sofia Mouchtaris**: Writing - Original draft preparation, Software, Validation. **Chetan Vadali**: Writing - Original draft preparation, Software, Validation. **Thomas Campbell Arnold**: Methodology, Writing - Review & Editing. **James J. Gugger**: Data Curation, Writing - Review & Editing. **Catherine Kulick**: Data Curation, Writing - Review & Editing. **Mariam Josyula**: Data Curation, Writing - Review & Editing. **Nina Petillo**: Data Curation, Writing - Review & Editing. **Sandhitsu Das**: Data Curation, Conceptualization, Writing - Review & Editing. **Jacob Dubroff**: Data Curation, Conceptualization, Writing - Review & Editing. **John A. Detre**: Data Curation, Conceptualization, Writing - Review & Editing. **Joel M. Stein**: Data Curation, Conceptualization, Methodology, Writing - Review & Editing. **Kathryn A. Davis**: Supervision, Project administration, Funding acquisition, Data Curation, Conceptualization, Writing - Review & Editing.

