## Supplementary Table for "Enhancing the Diagnostic Utility of ASL Imaging in Temporal Lobe Epilepsy through FlowGAN: An ASL to PET Image Translation Framework"

### Supplementary Methods

### Arterial Spin-Labeled MRI Preprocessing and Cerebral Blood Flow Computation

Arterial spin-labeled MRI images were preprocessed using *ASLPrep* 0.6.0 (Adebimpe et al. 2022, 2023), which is based on *fMRIPrep* (Esteban et al. (2019); Esteban et al. (2020); RRID:SCR_016216) and *Nipype* 1.8.6 (Gorgolewski et al. 2011).

### Anatomical data preprocessing

A total of 1 T1-weighted (T1w) images were found within the input BIDS dataset. The T1w image was corrected for intensity non-uniformity (INU) with N4BiasFieldCorrection (Tustison et al. 2010), distributed with ANTs 2.3.3 (Avants et al. 2008, RRID:SCR_004757), and used as T1w-reference throughout the workflow.

The T1w-reference was then skull-stripped with a *Nipype* implementation of the antsBrainExtraction.sh workflow (from ANTs), using OASIS30ANTs as target template. Brain tissue segmentation of cerebrospinal fluid (CSF), white-matter (WM) and gray-matter (GM) was performed on the brain-extracted T1w using fast (FSL 6.0.7.1, RRID:SCR_002823, Zhang, Brady, and Smith 2001). Brain surfaces were reconstructed using recon-all (FreeSurfer 6.0.1, RRID:SCR_001847, Dale, Fischl, and Sereno 1999), and the brain mask estimated previously was refined with a custom variation of the method to reconcile ANTs-derived and FreeSurfer-derived segmentations of the cortical gray-matter of Mindboggle (RRID:SCR_002438, Klein et al. 2017). Volume-based spatial normalization to one standard space (MNI152NLin2009cAsym) was performed through nonlinear registration with antsRegistration (ANTs 2.3.3), using brain-extracted versions of both T1w reference and the T1w template. The following template was selected for spatial normalization and accessed with *TemplateFlow* (23.1.0, Ciric et al. 2022): *ICBM 152 Nonlinear Asymmetrical template version 2009c* [Fonov et al. (2011), RRID:SCR_008796; TemplateFlow ID: MNI152NLin2009cAsym].

### ASL data preprocessing

For each of the 1 ASL runs found per subject (across all tasks and sessions), the following preprocessing was performed. First, a reference volume was generated using a custom methodology of *ASLPrep*, for use in head motion correction. Head-motion parameters were estimated for the ASL data using *FSL*'s mcflirt (Jenkinson et al. 2002). Motion correction was performed separately for each of the volume types to account for intensity differences between different contrasts, which, when motion corrected together, can conflate intensity differences with head motions (Wang et al. 2008). Next, ASLPrep concatenated the motion parameters across volume types and re-calculated relative root mean-squared deviation. The reference was then co-registered to the T1w reference using bbregister (FreeSurfer) which implements boundary-based registration (Greve and Fischl 2009). Co-registration was configured with six degrees of freedom.

#### *Cerebral blood flow computation and denoising*

*ASLPrep* calculated cerebral blood flow (CBF) from the single-delay PCASL using a single-compartment general kinetic model (Buxton et al. 1998). Calibration (M0) volumes associated with the ASL scan were smoothed with a Gaussian kernel (FWHM=5 mm) and the average calibration image was calculated and scaled by 10.0.

**Copyright Waiver**
The above methods description was automatically generated by *ASLPrep* with the express intention that users should copy and paste this text into their manuscripts unchanged. It is released under the unchanged CC0 license.

Dolui, Sudipto, Ze Wang, Russell T. Shinohara, David A. Wolk, John A. Detre, and for the Alzheimer’s Disease Neuroimaging Initiative. 2017. “Structural Correlation-Based Outlier Rejection (SCORE) Algorithm for Arterial Spin Labeling Time Series: SCORE: Denoising

Algorithm for ASL." *Journal of Magnetic Resonance Imaging* 45 (6): 1786–97. https://doi.org/10.1002/jmri.25436.

Dolui, Sudipto, Ronald Wolf, Seyed Ali Nabavizadeh, David A. Wolk, and John A. Detre. 2016. "Automated Quality Evaluation Index for 2D Asl Cbf Maps." *International Society for Magnetic Resonance in Medicine*, no. 1. https://doi.org/http://indexsmart.mirasmart.com/ISMRM2017/PDFfiles/0682.html.

Dolui, Sudipto, David A. Wolk, David A. Wolk, and John A. Detre. 2016. "SCRUB: A Structural Correlation and Empirical Robust Bayesian Method for Asl Data." *International Society for Magnetic Resonance in Medicine*, no. 1. https://doi.org/http://archive.ismrm.org/2016/2880.html.

Esteban, Oscar, Rastko Ciric, Karolina Finc, Ross W Blair, Christopher J Markiewicz, Craig A Moodie, James D Kent, et al. 2020. "Analysis of Task-Based Functional Mri Data Preprocessed with fMRIPrep." *Nature Protocols* 15 (7). Nature Publishing Group: 2186–2202. https://doi.org/10.1038/s41596-020-0327-3.

Esteban, Oscar, Christopher J Markiewicz, Ross W Blair, Craig A Moodie, A Ilkay Isik, Asier Erramuzpe, James D Kent, et al. 2019. "FMRIPrep: A Robust Preprocessing Pipeline for Functional Mri." *Nature Methods* 16 (1). Nature Publishing Group: 111–16. https://doi.org/10.1038/s41592-018-0235-4.

Fonov, Vladimir, Alan C. Evans, Kelly Botteron, C. Robert Almli, Robert C. McKinstry, and D. Louis Collins. 2011. "Unbiased Average Age-Appropriate Atlases for Pediatric Studies." *NeuroImage* 54 (1): 313–27. https://doi.org/10.1016/j.neuroimage.2010.07.033.

Glasser, Matthew F., Timothy S. Coalson, Emma C. Robinson, Carl D. Hacker, John Harwell, Essa Yacoub, Kamil Ugurbil, et al. 2016. "A Multi-Modal Parcellation of Human Cerebral Cortex." *Nature* 536 (7615): 171–78. https://doi.org/10.1038/nature18933.

Glasser, Matthew F., Stamatios N. Sotiropoulos, J. Anthony Wilson, Timothy S. Coalson, Bruce Fischl, Jesper L. Andersson, Junqian Xu, et al. 2013. "The Minimal Preprocessing Pipelines for the Human Connectome Project." *NeuroImage* 80 (October): 105–24. https://doi.org/10.1016/j.neuroimage.2013.04.127.

Gordon, Evan M., Timothy O. Laumann, Babatunde Adeyemo, Jeremy F. Huckins, William M. Kelley, and Steven E. Petersen. 2016. "Generation and Evaluation of a Cortical Area Parcellation from Resting-State Correlations." *Cerebral Cortex* 26 (1): 288–303. https://doi.org/10.1093/cercor/bhu239.

Gorgolewski, Krzysztof, Christopher D. Burns, Cindee Madison, Dav Clark, Yaroslav O. Halchenko, Michael L. Waskom, and Satrajit S. Ghosh. 2011. "Nipype: A Flexible, Lightweight and Extensible Neuroimaging Data Processing Framework in Python." *Frontiers in Neuroinformatics* 5. https://doi.org/10.3389/fninf.2011.00013.

Greve, Douglas N., and Bruce Fischl. 2009. "Accurate and Robust Brain Image Alignment Using Boundary-Based Registration." *NeuroImage* 48 (1): 63–72. https://doi.org/10.1016/j.neuroimage.2009.06.060.

Harris, Charles R., Jarrod K. Millman, Stéfan J. van der Walt, Ralf Gommers, Pauli Virtanen, David Cournapeau, Eric Wieser, et al. 2020. "Array Programming with NumPy." *Nature* 585 (7825): 357–62. https://doi.org/10.1038/s41586-020-2649-2.

Jenkinson, Mark, Peter Bannister, Michael Brady, and Stephen Smith. 2002. "Improved Optimization for the Robust and Accurate Linear Registration and Motion Correction of Brain Images." *NeuroImage* 17 (2): 825–41. https://doi.org/10.1016/s1053-8119(02)91132-8.

King, Maedbh, Carlos R Hernandez-Castillo, Russell A Poldrack, Richard B Ivry, and Jörn Diedrichsen. 2019. "Functional Boundaries in the Human Cerebellum Revealed by a Multi-Domain Task Battery." *Nature Neuroscience* 22 (8). Nature Publishing Group US New York: 1371–8. https://doi.org/10.1038/s41593-019-0436-x.

Klein, Arno, Satrajit S. Ghosh, Forrest S. Bao, Joachim Giard, Yrjö Häme, Eliezer Stavsky, Noah Lee, et al. 2017. "Mindboggling Morphometry of Human Brains." *PLOS Computational Biology* 13 (2): e1005350. https://doi.org/10.1371/journal.pcbi.1005350.

Lanczos, C. 1964. "Evaluation of Noisy Data." *Journal of the Society for Industrial and Applied Mathematics Series B Numerical Analysis* 1 (1): 76–85. https://doi.org/10.1137/0701007.

Najdenovska, Elena, Yasser Alemán-Gómez, Giovanni Battistella, Maxime Descoteaux, Patric Hagmann, Sebastien Jacquemont, Philippe Maeder, Jean-Philippe Thiran, Eleonora Fornari, and Meritxell Bach Cuadra. 2018. "In-Vivo Probabilistic Atlas of Human Thalamic Nuclei Based on Diffusion-Weighted Magnetic Resonance Imaging." *Scientific Data* 5 (1). Nature Publishing Group: 1–11. https://doi.org/10.1038/sdata.2018.270.

Pauli, Wolfgang M, Amanda N Nili, and J Michael Tyszka. 2018. "A High-Resolution Probabilistic in Vivo Atlas of Human Subcortical Brain Nuclei." *Scientific Data* 5 (1). Nature Publishing Group: 1–13. https://doi.org/10.1038/sdata.2018.63.

Power, Jonathan D., Anish Mitra, Timothy O. Laumann, Abraham Z. Snyder, Bradley L. Schlaggar, and Steven E. Petersen. 2014. "Methods to Detect, Characterize, and Remove Motion Artifact in Resting State fMRI." *NeuroImage* 84 (January): 320–41. https://doi.org/10.1016/j.neuroimage.2013.08.048.

Schaefer, Alexander, Ru Kong, Evan M. Gordon, Timothy O. Laumann, Xi-Nian Zuo, Avram J. Holmes, Simon B. Eickhoff, and B. T. Thomas Yeo. 2018. "Local-Global Parcellation of the Human Cerebral Cortex from Intrinsic Functional Connectivity

MRI." *Cerebral Cortex (New York, N.Y.: 1991)* 28 (9): 3095–3114. https://doi.org/10.1093/cercor/bhx179.

Tian, Ye, Daniel S Margulies, Michael Breakspear, and Andrew Zalesky. 2020. "Topographic Organization of the Human Subcortex Unveiled with Functional Connectivity Gradients." *Nature Neuroscience* 23 (11). Nature Publishing Group: 1421–32. https://doi.org/10.1038/s41593-020-00711-6.

Tustison, Nicholas J., Brian B. Avants, Philip A. Cook, Yuanjie Zheng, Alexander Egan, Paul A. Yushkevich, and James C. Gee. 2010. "N4ITK: Improved N3 Bias Correction." *IEEE Transactions on Medical Imaging* 29 (6): 1310–20. https://doi.org/10.1109/TMI.2010.2046908.

Virtanen, Pauli, Ralf Gommers, Travis E. Oliphant, Matt Haberland, Tyler Reddy, David Cournapeau, Evgeni Burovski, et al. 2020. "SciPy 1.0: Fundamental Algorithms for Scientific Computing in Python." *Nature Methods* 17 (3): 261–72. https://doi.org/10.1038/s41592-019-0686-2.

Wang, Ze, Geoffrey K Aguirre, Hengyi Rao, Jiongjiong Wang, María A Fernández-Seara, Anna R Childress, and John A Detre. 2008. "Empirical Optimization of Asl Data Analysis Using an Asl Data Processing Toolbox: ASLtbx." *Magnetic Resonance Imaging* 26 (2). Elsevier: 261–69. https://doi.org/10.1016/j.mri.2007.07.003.

Zhang, Y., M. Brady, and S. Smith. 2001. "Segmentation of Brain MR Images Through a Hidden Markov Random Field Model and the Expectation-Maximization Algorithm." *IEEE Transactions on Medical Imaging* 20 (1): 45–57. https://doi.org/10.1109/42.906424.

**Supplementary Table 1:** Correlation between PET SUV and ASL CBF values across brain regions

| DKT Atlas Region | Laterality | PET-ASL Spearman Correlation | p-value | Corrected p-value |
|---|---|---|---|---|
| Left-Hippocampus | Left | -0.139710654 | 0.255828 | 0.381989517 |
| CC_Posterior | Center | -0.113600794 | 0.35632 | 0.473645437 |
| ctx-lh-transversetemporal | Left | -0.081536054 | 0.508612 | 0.629985773 |
| ctx-lh-unknown | Left | -0.043325572 | 0.725729 | 0.816758922 |
| Right-Putamen | Right | -0.037141657 | 0.763641 | 0.832368353 |
| Left-Putamen | Left | -0.037141657 | 0.763641 | 0.832368353 |
| Right-Pallidum | Right | -0.023972211 | 0.846142 | 0.896200589 |
| Left-Amygdala | Left | -0.018170019 | 0.883078 | 0.924227114 |
| Left-Cerebellum-Cortex | Left | -0.002672062 | 0.982746 | 0.982746371 |
| Right-Cerebral-White-Matter | Right | 0.007558117 | 0.951223 | 0.960030506 |
| Left-Cerebellum-White-Matter | Left | 0.010230179 | 0.934012 | 0.951470062 |
| Left-Inf-Lat-Vent | Left | 0.014734512 | 0.905071 | 0.930686441 |
| CC_Anterior | Center | 0.023857694 | 0.846868 | 0.896200589 |
| Left-Cerebral-White-Matter | Left | 0.032217429 | 0.794236 | 0.857145801 |
| Left-Thalamus | Left | 0.041913196 | 0.734334 | 0.816758922 |
| Right-Cerebellum-White-Matter | Right | 0.04313471 | 0.72689 | 0.816758922 |
| Right-Amygdala | Right | 0.044356224 | 0.719471 | 0.816758922 |
| ctx-lh-superiortemporal | Left | 0.048287972 | 0.695769 | 0.806795591 |
| ctx-rh-unknown | Right | 0.050120243 | 0.68482 | 0.802638353 |
| Right-Inf-Lat-Vent | Right | 0.064816582 | 0.59949 | 0.710264894 |
| ctx-lh-insula | Left | 0.073558041 | 0.551083 | 0.660088619 |
| Right-Cerebellum-Cortex | Right | 0.078215063 | 0.526086 | 0.644307214 |
| ctx-lh-entorhinal | Left | 0.081879605 | 0.506822 | 0.629985773 |
| Right-Hippocampus | Right | 0.085887697 | 0.486175 | 0.616198252 |
| Right-vessel | Right | 0.092657343 | 0.462879 | 0.593574534 |
| ctx-rh-entorhinal | Right | 0.099858762 | 0.417815 | 0.542164775 |
| ctx-lh-parahippocampal | Left | 0.100049624 | 0.416921 | 0.542164775 |
| ctx-lh-inferiortemporal | Left | 0.117570714 | 0.339651 | 0.457061046 |
| ctx-rh-parsorbitalis | Right | 0.128373478 | 0.296813 | 0.404407245 |
| Left-Accumbens-area | Left | 0.130969195 | 0.28707 | 0.396084539 |
| Left-choroid-plexus | Left | 0.131083712 | 0.286646 | 0.396084539 |
| Right-VentralDC | Right | 0.132419743 | 0.281719 | 0.396084539 |
| ctx-lh-medialorbitofrontal | Left | 0.13337405 | 0.278235 | 0.396084539 |
| ctx-rh-lateraloccipital | Right | 0.134900943 | 0.27272 | 0.396084539 |
| ctx-lh-postcentral | Left | 0.152345688 | 0.214887 | 0.329897564 |
| ctx-lh-lateraloccipital | Left | 0.152422033 | 0.214655 | 0.329897564 |
| CC_Central | Center | 0.154406993 | 0.208673 | 0.329643345 |

| | | | | |
|---|---|---|---|---|
| ctx-rh-precentral | Right | 0.155055922 | 0.206744 | 0.329643345 |
| Right-Thalamus | Right | 0.156315609 | 0.203034 | 0.329643345 |
| ctx-rh-fusiform | Right | 0.157040883 | 0.20092 | 0.329643345 |
| ctx-lh-precentral | Left | 0.157957018 | 0.198272 | 0.329643345 |
| ctx-lh-fusiform | Left | 0.15849143 | 0.196738 | 0.329643345 |
| ctx-rh-lingual | Right | 0.165362446 | 0.177777 | 0.312543584 |
| ctx-lh-parsorbitalis | Left | 0.169752262 | 0.166381 | 0.297302946 |
| ctx-rh-transversetemporal | Right | 0.17303508 | 0.158215 | 0.287424747 |
| ctx-rh-lateralorbitofrontal | Right | 0.173111425 | 0.158029 | 0.287424747 |
| ctx-rh-postcentral | Right | 0.176928656 | 0.14892 | 0.279867245 |
| ctx-lh-lateralorbitofrontal | Left | 0.181967401 | 0.137504 | 0.262945364 |
| ctx-rh-superiortemporal | Right | 0.182081918 | 0.137252 | 0.262945364 |
| CC_Mid_Anterior | Center | 0.188991106 | 0.122706 | 0.243181603 |
| ctx-lh-lingual | Left | 0.190594343 | 0.119506 | 0.241224559 |
| Left-Pallidum | Left | 0.193571783 | 0.113732 | 0.233901373 |
| ctx-rh-insula | Right | 0.195633088 | 0.109862 | 0.230287654 |
| ctx-lh-parstriangularis | Left | 0.197541703 | 0.10637 | 0.230287654 |
| Left-VentralDC | Left | 0.19815246 | 0.105271 | 0.230287654 |
| ctx-lh-caudalanteriorcingulate | Left | 0.20078635 | 0.100632 | 0.228518852 |
| ctx-lh-paracentral | Left | 0.220941329 | 0.070203 | 0.162811581 |
| ctx-rh-parahippocampal | Right | 0.222315532 | 0.068432 | 0.162155047 |
| ctx-lh-rostralmiddlefrontal | Left | 0.224300492 | 0.065938 | 0.159716888 |
| ctx-lh-middletemporal | Left | 0.226705348 | 0.063015 | 0.156105151 |
| ctx-lh-superiorfrontal | Left | 0.227850517 | 0.06166 | 0.156105151 |
| Right-choroid-plexus | Right | 0.232125816 | 0.056809 | 0.147433209 |
| ctx-lh-cuneus | Left | 0.238844142 | 0.049817 | 0.132439602 |
| ctx-lh-posteriorcingulate | Left | 0.243462992 | 0.045431 | 0.123800181 |
| ctx-rh-supramarginal | Right | 0.250677558 | 0.039221 | 0.109617109 |
| ctx-rh-posteriorcingulate | Right | 0.261633011 | 0.031148 | 0.091760802 |
| ctx-rh-medialorbitofrontal | Right | 0.262663664 | 0.030466 | 0.091760802 |
| ctx-rh-rostralanteriorcingulate | Right | 0.265679276 | 0.028543 | 0.088889802 |
| CC_Mid_Posterior | Center | 0.26888575 | 0.02661 | 0.085309646 |
| ctx-rh-isthmuscingulate | Right | 0.274344391 | 0.023575 | 0.077868444 |
| ctx-lh-parsopercularis | Left | 0.274420735 | 0.023535 | 0.077868444 |
| ctx-rh-inferiorparietal | Right | 0.274611597 | 0.023434 | 0.077868444 |
| Left-Caudate | Left | 0.286139634 | 0.018005 | 0.065418489 |
| ctx-lh-caudalmiddlefrontal | Left | 0.288964385 | 0.016853 | 0.065418489 |
| ctx-lh-isthmuscingulate | Left | 0.294041302 | 0.01494 | 0.060311766 |
| **ctx-lh-supramarginal** | **Left** | **0.305378478** | **0.011331** | **0.047501315** |
| **ctx-rh-caudalanteriorcingulate** | **Right** | **0.318204375** | **0.008182** | **0.035675072** |
| **ctx-rh-superiorparietal** | **Right** | **0.319349544** | **0.007943** | **0.035675072** |
| **ctx-rh-superiorfrontal** | **Right** | **0.320838264** | **0.00764** | **0.035675072** |
| **Right-Accumbens-area** | **Right** | **0.32209795** | **0.007392** | **0.035675072** |
| **ctx-rh-cuneus** | **Right** | **0.326640455** | **0.006556** | **0.034026878** |

| | | | | |
|---|---|---|---|---|
| ctx-lh-precuneus | Left | 0.329236172 | 0.006116 | 0.033331804 |
| ctx-rh-inferiortemporal | Right | 0.32969424 | 0.006041 | 0.033331804 |
| ctx-lh-rostralanteriorcingulate | Left | 0.343894339 | 0.004087 | 0.024747733 |
| ctx-rh-middletemporal | Right | 0.345383059 | 0.003919 | 0.024747733 |
| ctx-rh-pericalcarine | Right | 0.345955644 | 0.003856 | 0.024747733 |
| ctx-lh-inferiorparietal | Left | 0.347826087 | 0.003656 | 0.024747733 |
| ctx-rh-paracentral | Right | 0.351070733 | 0.003331 | 0.024747733 |
| ctx-lh-pericalcarine | Left | 0.364545559 | 0.002241 | 0.018788632 |
| ctx-rh-parstriangularis | Right | 0.370309577 | 0.001881 | 0.017988409 |
| Right-Caudate | Right | 0.38275375 | 0.001276 | 0.015451327 |
| ctx-rh-caudalmiddlefrontal | Right | 0.417414208 | 0.000398 | 0.006384605 |
| ctx-rh-parsopercularis | Right | 0.422987365 | 0.000326 | 0.006384605 |
| ctx-rh-precuneus | Right | 0.439363286 | 0.000178 | 0.004848686 |
| ctx-rh-rostralmiddlefrontal | Right | 0.455815551 | 9.38E-05 | 0.003407828 |
| ctx-lh-superiorparietal | Left | 0.501584151 | 1.32E-05 | 0.000717685 |

**Supplementary Table 2:** Correlation between PET SUV and ASL CBF asymmetry values across brain regions

| Region Name | Spearman Correlation | p-value | corrected p-value |
|---|---|---|---|
| pericalcarine | -0.085971686 | 0.6180962 | 0.672634123 |
| precuneus | -0.074646075 | 0.6652512 | 0.703265505 |
| Putamen | 0.010810811 | 0.950103 | 0.950103021 |
| parahippocampal | 0.022393822 | 0.8968535 | 0.921766125 |
| lateralorbitofrontal | 0.114543115 | 0.5059233 | 0.567247367 |
| Pallidum | 0.185585586 | 0.2785139 | 0.322031677 |
| transversetemporal | 0.203861004 | 0.2330371 | 0.278141017 |
| entorhinal | 0.204890605 | 0.2306345 | 0.278141017 |
| parsorbitalis | 0.23011583 | 0.1769618 | 0.22577883 |
| Amygdala | 0.263577864 | 0.1203596 | 0.159046626 |
| supramarginal | 0.275675676 | 0.1036544 | 0.142044931 |
| Caudate | 0.281081081 | 0.0967862 | 0.137734151 |
| fusiform | 0.281338481 | 0.096468 | 0.137734151 |
| Hippocampus | 0.296782497 | 0.0787993 | 0.121482321 |
| insula | 0.313513514 | 0.0626106 | 0.100721368 |
| parsopercularis | 0.315315315 | 0.061037 | 0.100721368 |
| postcentral | 0.321750322 | 0.0556707 | 0.098086416 |
| inferiorparietal | 0.334620335 | 0.0460661 | 0.085222286 |
| rostralmiddlefrontal | 0.352638353 | 0.0349053 | 0.067973535 |
| **precentral** | **0.393822394** | **0.0174788** | **0.035928708** |
| **isthmuscingulate** | **0.404890605** | **0.0143043** | **0.031132844** |
| **lingual** | **0.406692407** | **0.0138366** | **0.031132844** |
| **superiorfrontal** | **0.407979408** | **0.0135105** | **0.031132844** |
| **posteriorcingulate** | **0.412097812** | **0.0125101** | **0.031132844** |
| **Thalamus** | **0.432432432** | **0.0084394** | **0.024019835** |
| **cuneus** | **0.433719434** | **0.0082252** | **0.024019835** |
| **paracentral** | **0.453539254** | **0.0054678** | **0.018391755** |
| **superiortemporal** | **0.457400257** | **0.0050356** | **0.018391755** |
| **caudalmiddlefrontal** | **0.47001287** | **0.0038227** | **0.015715511** |
| **rostralanteriorcingulate** | **0.479279279** | **0.0031014** | **0.014343828** |
| **superiorparietal** | **0.516602317** | **0.0012567** | **0.006642763** |
| **parstriangularis** | **0.523552124** | **0.00105** | **0.006474949** |
| **caudalanteriorcingulate** | **0.542599743** | **0.0006288** | **0.004653126** |
| **inferiortemporal** | **0.568854569** | **0.0002945** | **0.002724049** |
| **middletemporal** | **0.571685972** | **0.0002703** | **0.002724049** |
| **medialorbitofrontal** | **0.578893179** | **0.0002166** | **0.002724049** |
| **lateraloccipital** | **0.64041184** | **2.591E-05** | **0.000958699** |

**Supplementary Table 3:** Descriptive AUC values for lateralizing ability for PET SUV asymmetry and ASL CBF asymmetry

| Region Name | AUC PET | AUC PET CI | AUC ASL | AUC ASL CI | AUC Difference | DeLong p-value | DeLong p-value - corrected |
|---|---|---|---|---|---|---|---|
| **Amygdala** | **0.8695652** | **(0.7339684599239622, 1)** | **0.5451505** | **(0.34548447251407666, 0.7448165308304049)** | **0.324414716** | **0.000526498** | **0.019480435** |
| fusiform | 0.9130435 | (0.8003039255257205, 1) | 0.67892977 | (0.4894965487628896, 0.8683629830096856) | 0.234113712 | 0.0314008 | 0.387276531 |
| Hippocampus | 0.9297659 | (0.8278206136277839, 1) | 0.69899666 | (0.5127814167226135, 0.8852118943141758) | 0.230769231 | 0.023242128 | 0.387276531 |
| Thalamus | 0.8963211 | (0.7740281429882835, 1) | 0.69899666 | (0.5127814167226135, 0.8852118943141758) | 0.197324415 | 0.063993633 | 0.417964881 |
| parahippocampal | 0.7892977 | (0.6239633573873283, 0.9546319603384242) | 0.60869565 | (0.41155861684967737, 0.8058326874981487) | 0.180602007 | 0.157089935 | 0.604091343 |
| insula | 0.8963211 | (0.7740281429882834, 1) | 0.72575251 | (0.5445861697200628, 0.9069188470023454) | 0.170568562 | 0.114276747 | 0.528529953 |
| inferiortemporal | 0.9698997 | (0.9023008693544359, 1) | 0.82943144 | (0.6773575698601361, 0.9815053063940445) | 0.140468227 | 0.067778089 | 0.417964881 |
| entorhinal | 0.8461538 | (0.7005056304775892, 0.9918020618301033) | 0.71237458 | (0.528572638523587, 0.8961765253560116) | 0.133779264 | 0.262238041 | 0.693057679 |
| paracentral | 0.541806 | (0.34211848715483556, 0.7414935529789437) | 0.4180602 | (0.22519382790107645, 0.6109265734367161) | 0.123745819 | 0.250048165 | 0.693057679 |
| parsopercularis | 0.7993311 | (0.637049612403164, 0.9616125949546955) | 0.68561873 | (0.4972056050619762, 0.8740318531320039) | 0.113712375 | 0.347589259 | 0.756517798 |
| precuneus | 0.6923077 | (0.5049669824973455, 0.8796484021180391) | 0.57859532 | (0.3797533523222573, 0.7774372831292478) | 0.113712375 | 0.47330388 | 0.890823133 |
| superiortemporal | 0.9397993 | (0.8450892007486996, 1) | 0.83277592 | (0.681941135406288, 0.9836107040585947) | 0.107023411 | 0.229337248 | 0.693057679 |
| medialorbitofrontal | 0.7525084 | (0.5773126826777769, 0.9277040397302497) | 0.65886288 | (0.46667659510579074, 0.8510491574025705) | 0.093645485 | 0.329099742 | 0.756517798 |
| middletemporal | 0.9464883 | (0.8569970213439179, 1) | 0.85618729 | (0.7146859196711209, 0.9976886622686784) | 0.090301003 | 0.214648199 | 0.693057679 |
| transversetemporal | 0.8026756 | (0.6414488367377037, 0.9639023338308582) | 0.71237458 | (0.528572638523587, 0.8961765253560116) | 0.090301003 | 0.457808341 | 0.890823133 |
| posteriorcingulate | 0.7591973 | (0.5856469255733981, 0.9327477232560334) | 0.69565217 | (0.5088675197227595, 0.8824368281033277) | 0.063545151 | 0.549744766 | 0.924570744 |
| precentral | 0.7391304 | (0.5608298614076075, 0.9174310081576098) | 0.68561873 | (0.4972056050619762, 0.8740318531320039) | 0.053511706 | 0.624663255 | 0.941684159 |
| postcentral | 0.7190635 | (0.5365511488325039, 0.9015759414684994) | 0.67558528 | (0.48566142330010054, 0.8655091452617724) | 0.043478261 | 0.747003804 | 0.941684159 |
| caudalmiddlefrontal | 0.5117057 | (0.31231651078230227, 0.7110948604551559) | 0.47491639 | (0.27708347712490033, 0.6727492987948321) | 0.036789298 | 0.724870767 | 0.941684159 |
| parstriangularis | 0.819398 | (0.6637375070937672, 0.9750584795283064) | 0.78595318 | (0.6196374810897176, 0.9522688734253326) | 0.033444816 | 0.731724692 | 0.941684159 |
| Putamen | 0.5618729 | (0.362479869577453, 0.7612659498205405) | 0.52842809 | (0.3287641495610004, 0.7280920377299697) | 0.033444816 | 0.796524589 | 0.941684159 |
| inferiorparietal | 0.7926421 | (0.6283071577348349, 0.95697712320162) | 0.76588629 | (0.59404492859981, 0.937727646651026) | 0.026755853 | 0.832749507 | 0.941684159 |
| superiorfrontal | 0.6989967 | (0.5127814167226137, 0.8852118943141758) | 0.6722408 | (0.48183912209115076, 0.8626424832600198) | 0.026755853 | 0.806142045 | 0.941684159 |
| isthmuscingulate | 0.735786 | (0.5567468913344518, 0.9148250150200632) | 0.71237458 | (0.528572638523587, 0.8961765253560116) | 0.023411371 | 0.833953761 | 0.941684159 |
| supramarginal | 0.826087 | (0.6727960661644309, 0.9793778468790474) | 0.81270903 | (0.6547621860380669, 0.970655874162602) | 0.013377926 | 0.909596128 | 0.941684159 |
| Caudate | 0.5117057 | (0.3123165107823024, 0.711094860455156) | 0.50167224 | (0.3025780413037866, 0.7007664403015645) | 0.010033445 | 0.930259436 | 0.941684159 |
| Pallidum | 0.4983278 | (0.2993535000096256, 0.6973020183850233) | 0.5083612 | (0.3090595362785862, 0.7076628717481697) | -0.010033445 | 0.941684159 | 0.941684159 |
| lingual | 0.735786 | (0.5567468913344519, 0.9148250150200633) | 0.75250836 | (0.5773126826777771, 0.9277040397302497) | -0.016722408 | 0.870317044 | 0.941684159 |
| superiorparietal | 0.6120401 | (0.41515003673467554, 0.808930230823853) | 0.62876254 | (0.43328288051456715, 0.824242203097473) | -0.016722408 | 0.866750392 | 0.941684159 |
| rostralanteriorcingula | 0.5551839 | (0.35564852355594, 0.7547193694206488) | 0.58528428 | (0.3867414103551902, 0.7838271515177193) | -0.030100334 | 0.787336594 | 0.941684159 |
| lateralorbitofrontal | 0.7391304 | (0.5608298614076076, 0.9174310081576099) | 0.79598662 | (0.6326691497442802, 0.959304094402877) | -0.056856187 | 0.66542839 | 0.941684159 |
| cuneus | 0.4147157 | (0.22223856299378186, 0.6071928751333086) | 0.4916388 | (0.2929367842822041, 0.69034080769104) | -0.076923077 | 0.488408865 | 0.890823133 |
| rostralmiddlefrontal | 0.6287625 | (0.43328288051456715, 0.824242203097473) | 0.71237458 | (0.528572638523587, 0.8961765253560116) | -0.08361204 | 0.505602318 | 0.890823133 |
| caudalanteriorcingula | 0.6254181 | (0.42963271841336304, 0.8212034019879748) | 0.71571906 | (0.5325548905512634, 0.8988832365390376) | -0.090301003 | 0.346111387 | 0.756517798 |
| lateraloccipital | 0.7458194 | (0.5690407712564161, 0.9225980247302061) | 0.87625418 | (0.7437846308589233, 1) | -0.130434783 | 0.163267931 | 0.604091343 |
| parsorbitalis | 0.5050167 | (0.3058133834021711, 0.7042200614138824) | 0.72240803 | (0.5405615337576926, 0.904254519754013) | -0.217391304 | 0.089687817 | 0.474064173 |
| pericalcarine | 0.4548495 | (0.25841436303226284, 0.6512846336232556) | 0.73913043 | (0.5608298614076075, 0.9174310081576098) | -0.284280936 | 0.047200129 | 0.417964881 |

**Supplementary Table 4**: Asymmetry correlation values between original PET and ASL, original and synthetic PET, and their difference, for the test set subjects

| Region Name | Spearman PET-ASL | p-value PET-ASL | p-value PET-ASL - corrected | Spearman PET-synthetic PET | p-value PET-synthetic PET | p-value PET-synthetic PET - corrected | Correlation Difference (PET-ASL minus PET-synthetic PET) |
|---|---|---|---|---|---|---|---|
| medialorbitofrontal | 0.738596491 | 0.000304302 | 0.005045578 | 0.856140351 | 2.92054E-06 | 0.00010806 | 0.11754386 |
| cuneus | 0.398245614 | 0.091266564 | 0.225124192 | 0.814035088 | 2.225E-05 | 0.000411625 | 0.415789474 |
| superiorparietal | 0.801754386 | 3.66414E-05 | 0.001355732 | 0.763157895 | 0.000144179 | 0.001778209 | -0.038596491 |
| lateraloccipital | 0.728070175 | 0.000409101 | 0.005045578 | 0.719298246 | 0.000518354 | 0.004794777 | -0.00877193 |
| Pallidum | 0.235087719 | 0.332631898 | 0.49229521 | 0.643859649 | 0.002931293 | 0.021691567 | 0.40877193 |
| inferiorparietal | 0.447368421 | 0.054789329 | 0.150515209 | 0.610526316 | 0.005497326 | 0.029057297 | 0.163157895 |
| insula | 0.270175439 | 0.26327627 | 0.442782818 | 0.61754386 | 0.004843338 | 0.029057297 | 0.347368421 |
| middletemporal | 0.687719298 | 0.001137229 | 0.010519364 | 0.542105263 | 0.016496928 | 0.067820706 | -0.145614035 |
| precuneus | -0.045614035 | 0.852896074 | 0.940352299 | 0.549122807 | 0.014889998 | 0.067820706 | 0.594736842 |
| caudalmiddlefrontal | 0.5 | 0.029258036 | 0.105224349 | 0.528070175 | 0.020124316 | 0.074459968 | 0.028070175 |
| parsopercularis | 0.3 | 0.212074332 | 0.392337514 | 0.51754386 | 0.023237901 | 0.07686988 | 0.21754386 |
| isthmuscingulate | 0.277192982 | 0.250588895 | 0.441513767 | 0.512280702 | 0.024930772 | 0.07686988 | 0.235087719 |
| superiorfrontal | 0.443859649 | 0.056951701 | 0.150515209 | 0.484210526 | 0.035658766 | 0.08795829 | 0.040350877 |
| rostralmiddlefrontal | 0.389473684 | 0.099299102 | 0.229629172 | 0.484210526 | 0.035658766 | 0.08795829 | 0.094736842 |
| lingual | 0.115789474 | 0.636895929 | 0.785504979 | 0.485964912 | 0.034898352 | 0.08795829 | 0.370175439 |
| superiortemporal | 0.550877193 | 0.014508498 | 0.059646048 | 0.464912281 | 0.044896127 | 0.103822293 | -0.085964912 |
| paracentral | 0.338596491 | 0.156189745 | 0.339942386 | 0.449122807 | 0.053731735 | 0.115241181 | 0.110526316 |
| fusiform | 0.329824561 | 0.167887465 | 0.345102012 | 0.443859649 | 0.056951701 | 0.115241181 | 0.114035088 |
| supramarginal | -0.00877193 | 0.971568936 | 0.971568936 | 0.440350877 | 0.059177904 | 0.115241181 | 0.449122807 |
| postcentral | 0.494736842 | 0.031282914 | 0.105224349 | 0.424561404 | 0.070015189 | 0.1295281 | -0.070175439 |
| precentral | 0.224561404 | 0.35534599 | 0.505684678 | 0.407017544 | 0.083718971 | 0.140800088 | 0.18245614 |
| Hippocampus | 0.149122807 | 0.542322135 | 0.716639964 | 0.407017544 | 0.083718971 | 0.140800088 | 0.257894737 |
| transversetemporal | 0.150877193 | 0.53752721 | 0.716639964 | 0.347368421 | 0.145066057 | 0.233367135 | 0.196491228 |
| inferiortemporal | 0.668421053 | 0.001757332 | 0.013004254 | 0.336842105 | 0.158483043 | 0.244328025 | -0.331578947 |
| parstriangularis | 0.652631579 | 0.002454158 | 0.015133977 | 0.298245614 | 0.214890107 | 0.318034571 | -0.354385965 |
| Caudate | -0.042105263 | 0.864107518 | 0.940352299 | 0.292982456 | 0.223483753 | 0.318034571 | 0.335087719 |
| parahippocampal | -0.236842105 | 0.328931042 | 0.49229521 | 0.268421053 | 0.266509863 | 0.36521722 | 0.505263158 |
| posteriorcingulate | 0.456140351 | 0.049655396 | 0.150515209 | 0.196491228 | 0.420101534 | 0.55513417 | -0.259649123 |
| entorhinal | 0.240350877 | 0.321602437 | 0.49229521 | 0.185964912 | 0.445906275 | 0.560779561 | -0.054385965 |
| lateralorbitofrontal | 0.010526316 | 0.96588561 | 0.971568936 | 0.18245614 | 0.454686131 | 0.560779561 | 0.171929825 |
| pericalcarine | -0.126315789 | 0.606350247 | 0.773619281 | 0.161403509 | 0.509171343 | 0.607720635 | 0.287719298 |
| rostralanteriorcingulate | 0.301754386 | 0.20928288 | 0.392337514 | 0.136842105 | 0.57641738 | 0.666482595 | -0.164912281 |
| Thalamus | 0.628070175 | 0.003983054 | 0.021053285 | -0.114035088 | 0.642043574 | 0.719867037 | -0.742105263 |
| caudalanteriorcingulate | 0.619298246 | 0.004690234 | 0.021692334 | 0.064912281 | 0.791768213 | 0.861630115 | -0.554385965 |
| Putamen | 0.094736842 | 0.699657178 | 0.835074697 | 0.054385965 | 0.824989714 | 0.872131984 | -0.040350877 |
| parsorbitalis | 0.026315789 | 0.914837897 | 0.967114348 | -0.005263158 | 0.982939261 | 0.994312737 | -0.031578947 |
| Amygdala | -0.049122807 | 0.841711569 | 0.940352299 | 0.001754386 | 0.994312737 | 0.994312737 | 0.050877193 |

**Supplementary Table 5**: Congruency values between original PET and ASL, original and synthetic PET, and their difference, for the test set subjects

| Region Name | Congruency Original PET - ASL | Congruency Original PET - Synthetic PET | Congruency Difference |
|---|---|---|---|
| Hippocampus | 0.421052632 | 0.684210526 | 0.263157895 |
| Pallidum | 0.526315789 | 0.789473684 | 0.263157895 |
| inferiorparietal | 0.631578947 | 0.842105263 | 0.210526316 |
| medialorbitofrontal | 0.631578947 | 0.842105263 | 0.210526316 |
| insula | 0.578947368 | 0.789473684 | 0.210526316 |
| rostralanteriorcingulate | 0.473684211 | 0.631578947 | 0.157894737 |
| superiorfrontal | 0.526315789 | 0.684210526 | 0.157894737 |
| parahippocampal | 0.368421053 | 0.526315789 | 0.157894737 |
| lateralorbitofrontal | 0.526315789 | 0.684210526 | 0.157894737 |
| precentral | 0.684210526 | 0.842105263 | 0.157894737 |
| fusiform | 0.473684211 | 0.578947368 | 0.105263158 |
| Caudate | 0.526315789 | 0.631578947 | 0.105263158 |
| rostralmiddlefrontal | 0.789473684 | 0.894736842 | 0.105263158 |
| supramarginal | 0.631578947 | 0.736842105 | 0.105263158 |
| pericalcarine | 0.526315789 | 0.631578947 | 0.105263158 |
| isthmuscingulate | 0.631578947 | 0.684210526 | 0.052631579 |
| transversetemporal | 0.578947368 | 0.631578947 | 0.052631579 |
| precuneus | 0.578947368 | 0.631578947 | 0.052631579 |
| postcentral | 0.578947368 | 0.578947368 | 0 |
| Putamen | 0.473684211 | 0.473684211 | 0 |
| cuneus | 0.684210526 | 0.684210526 | 0 |
| middletemporal | 0.736842105 | 0.684210526 | -0.052631579 |
| parsopercularis | 0.736842105 | 0.684210526 | -0.052631579 |
| caudalmiddlefrontal | 0.631578947 | 0.578947368 | -0.052631579 |
| superiorparietal | 0.789473684 | 0.736842105 | -0.052631579 |
| lateraloccipital | 0.789473684 | 0.736842105 | -0.052631579 |
| Amygdala | 0.578947368 | 0.526315789 | -0.052631579 |
| posteriorcingulate | 0.684210526 | 0.631578947 | -0.052631579 |
| lingual | 0.684210526 | 0.631578947 | -0.052631579 |
| paracentral | 0.631578947 | 0.526315789 | -0.105263158 |
| entorhinal | 0.631578947 | 0.526315789 | -0.105263158 |
| superiortemporal | 0.736842105 | 0.578947368 | -0.157894737 |
| Thalamus | 0.631578947 | 0.473684211 | -0.157894737 |
| caudalanteriorcingulate | 0.736842105 | 0.526315789 | -0.210526316 |
| inferiortemporal | 0.789473684 | 0.526315789 | -0.263157895 |
| parsorbitalis | 0.631578947 | 0.368421053 | -0.263157895 |
| parstriangularis | 0.789473684 | 0.526315789 | -0.263157895 |
